# One size does not fit all. Genomics differentiates among binge-eating disorder, bulimia nervosa, and anorexia nervosa

**DOI:** 10.1101/2020.03.24.20042648

**Authors:** Christopher Hübel, Mohamed Abdulkadir, Moritz Herle, Ruth J.F. Loos, Gerome Breen, Cynthia M. Bulik, Nadia Micali

## Abstract

**Objective:** Genome-wide association studies have identified multiple genomic regions associated with anorexia nervosa. Relatively few or no genome-wide studies of other eating disorders, such as bulimia nervosa and binge-eating disorder, have been performed, despite their substantial heritability. Exploratively, we aimed to identify traits that are genetically associated with binge-type eating disorders.

**Method:** We calculated genome-wide polygenic scores for 269 trait and disease outcomes using PRSice v2.2 and their association with anorexia nervosa, bulimia nervosa, and binge-eating disorder in up to 640 cases and 17,050 controls from the UK Biobank. Significant associations were tested for replication in the Avon Longitudinal Study of Parents and Children (up to 217 cases and 3018 controls).

**Results:** Individuals with binge-type eating disorders had higher polygenic scores than controls for other psychiatric disorders, including depression, schizophrenia, and attention deficit hyperactivity disorder, and higher polygenic scores for body mass index.

**Discussion:** Our findings replicate some of the known comorbidities of eating disorders on a genomic level and motivate a deeper investigation of shared and unique genomic factors across the three primary eating disorders.

## Introduction

Eating disorders are complex and heritable psychiatric conditions (Yilmaz et al., 2015). The most studied is anorexia nervosa, which is characterised by dangerously low body weight and extreme fear of gaining weight, while engaging in behaviours that induce negative energy balance, such as fasting or, in some cases, excessive exercise (Treasure et al., 2015). In contrast, individuals with bulimia nervosa and binge-eating disorder experience episodes of excessive overeating (i.e., binge eating) accompanied by a sense of loss of control, in which they consume unusually large amounts of food in a short period of time. Individuals with bulimia nervosa also engage in recurrent compensatory behaviors (e.g., self-induced vomiting, laxative, or diuretic abuse) to counteract the effects of binge eating (American Psychiatric Association, 2013).

Twin, family, and adoption studies over the past 30 years have shown that eating disorders are heritable (Yilmaz et al., 2015). The largest genome-wide association study (GWAS) to date identified eight genomic regions associated with anorexia nervosa and also found genetic correlations with obsessive-compulsive disorder, schizophrenia, anxiety, and major depressive disorder. This implies that anorexia nervosa shares genetic risk variants with these phenotypes. However, the study also found that anorexia nervosa has significant negative genetic correlations with anthropometric traits, including BMI, fat mass, and fat-free mass. This means that anorexia nervosa risk variants are enriched for variants associated with lower BMI, fat mass, and fat-free mass. (Duncan et al., 2017; Hübel et al., 2019; Watson et al., 2019). The GWAS additionally highlighted a shared genetic basis between anorexia nervosa and high-density lipoprotein cholesterol, fasting insulin, as well as insulin sensitivity, suggesting that anorexia nervosa may have a metabolic component. It is unclear, however, if binge-type eating disorders may also have a metabolic component.

Although twin and family studies demonstrate a positive genetic correlation (∼0.46) between anorexia nervosa and bulimia nervosa, indicating considerable shared genetics (Bulik et al., 2010; Yao, Larsson, et al., 2019), GWASs of bulimia nervosa or binge-eating disorder with sufficient size have yet to be conducted to allow well powered investigations of their molecular genetic correlations with each other. On a phenotypic level, binge-eating disorder co-occurs with anxiety, depression, as well as attention deficit hyperactivity disorder, substance abuse, and personality disorders. In addition, individuals with binge-eating disorder commonly are overweight or obese (Hilbert, 2019). We have previously suggested that increased genetic liability for obesity was associated with increased risk of binge eating in adolescence (Abdulkadir et al., 2020). In contrast, it is not well understood the extent to which binge-type eating disorders share genetics with other psychiatric, anthropometric, and metabolic traits.

To explore differences in the genetics of eating disorders and generate new hypotheses regarding their genetic overlap with other traits, we conducted polygenic score analyses to identify traits that are genetically associated with either of the three primary eating disorders: anorexia nervosa, bulimia nervosa, or binge-eating disorder, in a subsample of the UK Biobank (Sudlow et al., 2015) with attempted replication in the Avon Longitudinal Study of Parents and Children (ALSPAC) cohort.

## Methods

### Discovery sample of the UK Biobank

The UK Biobank (ukbiobank.ac.uk) is a unique epidemiological resource to improve prevention, diagnosis, and treatment of psychiatric and somatic illnesses. The UK Biobank recruited participants from the general population between 2006–2010. All participants were between 40 to 69 years old, were registered with a general practitioner through the United Kingdom’s National Health Service, and lived within traveling distance of one of the assessment centers. The UK Biobank is approved by the North West Multi-centre Research Ethics Committee. Genomewide array data for this study were available for 488,363 individuals. All participants gave written consent. We identified non-European participants by 4-means clustering on the first two principal components derived from the genotype data, and excluded related individuals (KING relatedness metric >0.088, equivalent to a relatedness value of 0.25; *N* = 7,765).

We identified 1,488 participants who either self-reported any of the three primary eating disorders in the mental health questionnaire (Davis et al., 2020) or had an International Classification of Diseases, version 10 (ICD-10) (World Health Organization, 1992) hospital diagnosis of F50.0 or F50.1 for anorexia nervosa, or F50.2 or F50.3 for bulimia nervosa. This resulted in 768 (4.7%) participants with anorexia nervosa, 423 (2.7%) with bulimia nervosa, and 561 (3.5%) with binge-eating disorder. If participants self-reported more than one diagnosis, they were assigned to all groups. We randomly sampled one set of controls in the ratio of 1 to 10, resulting in 15,500 controls by using the following exclusion criteria: Controls must have answered the mental health questionnaire, were not diagnosed with a psychiatric disorder (i.e., self-report or ICD-10 diagnosis), or taking any psychotropic medication. The final analysis included 17,050 (92% female) European participants (**Supplementary Table 1**) representing 3.4% of the genotyped UK Biobank participants (n = 502,682). This study was completed under the UK Biobank approved study application 27546.

### Sample from the Avon Longitudinal Study of Parents and Children (ALSPAC)

The ALSPAC is a population-based sample of pregnant women and their children based in the former county of Avon, UK (Boyd et al., 2013; Fraser et al., 2013; Golding et al., 2001). Women expected to deliver from April 1, 1991, until December 31, 1992, were invited to participate. Children from 14,541 pregnancies were enrolled and 13,988 alive at 1 year. Additionally, 913 children were enrolled at age 7 years. All women gave written informed consent. Children at age 14 (wave 14, *n* = 10,581), 16 (wave 16, *n* = 9,702), and 18 years (wave 18, *n* = 9,505) that had not withdrawn consent were followed up. Parents answered questionnaires on 7,025 adolescents at wave 14 and on 5,656 at wave 16. With 6,140 (58%) responding at wave 14, 5,069 (52%) at wave 16, and 3,228 (34%) at wave 18 which was used to augment the validity of the probable eating disorder diagnoses (Micali et al., 2015). The study website contains details of all the data that is available through a fully searchable data dictionary and variable search tool (www.bristol.ac.uk/alspac/researchers/our-data/). To reduce potential confounding through genetic relatedness in our analyses, we removed randomly one individual of each pair that is closely related (*φ* > 0.2) using PLINK v1.90 (Chang et al., 2015), excluding 75 individuals that were duplicates, monozygotic twins, first-degree relatives (i.e., parent-offspring and full siblings), or second-degree relatives (i.e., half-siblings, uncles, aunts, grandparents, and double cousins, **Supplementary Table 2**).

We derived probable eating disorder diagnoses as previously reported (**Supplementary Table 3**) (Micali et al., 2015), using a combination of adolescent self-report as well as information on the adolescents provided by their parents, a gold standard for childhood psychiatric disorders.

Ethical approval for the ALSPAC participants of this study was obtained from the ALSPAC Ethics and Law Committee and the Local Research Ethics Committees: www.bristol.ac.uk/alspac/researchers/research-ethics/. Consent for biological samples has been collected in accordance with the Human Tissue Act (2004) and informed consent for the use of data collected via questionnaires and clinics was obtained from participants following the recommendations of the ALSPAC Ethics and Law Committee at the time.

### Polygenic risk scoring on eating disorder phenotypes in UK Biobank and replication in ALSPAC

We used PRSice (Choi & O’Reilly, 2019), version 2.2.3. We clumped the single nucleotide polymorphisms (SNPs) that were present both in the summary statistics of the trait and in the genotype data of the UK Biobank (i.e., overlapping SNPs) to obtain genetically independent SNPs. We retained the SNP with the smallest p value in each 250 kilobase window of all those in linkage disequilibrium (*r*^*2*^> 0.1). We calculated 269 polygenic scores at their optimal *p* value threshold in each individual by scoring the number of effect alleles (weighted by the allele effect size) across the set of remaining SNPs (for a full list, see **Supplementary Table 4**). We calculated the polygenic score using the high-resolution scoring (i.e., incrementally across a large number of *p* value thresholds) method to identify the *p* value threshold at which the polygenic score is optimally associated with the outcome and explains the most variance (i.e., resulting in the highest adjusted *R*^*2*^ for continuous outcomes and Nagelkerke’s *R*^*2*^ on the liability scale for binary outcomes). We evaluated the associations between polygenic score and eating disorder diagnosis using logistic regressions, adjusted for sex and the first six principal components that were calculated on the European subsample. To adjust for overfitting, we permuted case-control status at every *p* value threshold 10,000 times and, hence, calculated empirical *p* values. We converted the observed *R*^*2*^ to the liability scale assuming following population prevalences of 3% for anorexia nervosa, 1.8% for bulimia nervosa, and 3% for binge-eating disorder (Micali et al., 2017; Smink et al., 2014).

To correct for multiple testing (i.e., 269 polygenic score regression models), we calculated *Q* values using the false discovery rate approach (Benjamini & Hochberg, 1995; Benjamini & Yekutieli, 2001). We did not stratify analyses by sex because of the low number of male eating disorder cases, but included sex as a covariate. We used the UK Biobank sample as our discovery cohort and the SNPs that were included in the strongest associated polygenic score were used to derive polygenic scores in the ALSPAC sample, in which we repeated the analysis also adjusting for sex, and the first six ancestry-informative principal components.

## Results

Descriptive statistics of the analyses samples included from UK Biobank and ALSPAC are listed in Supplementary Tables 1 and 2.

### Polygenic scoring on eating disorder phenotypes in UK Biobank and ALSPAC

After correcting for multiple testing using the false discovery rate adjustment, 18 polygenic scores were significantly associated with the three primary eating disorders in the subsample of the UK Biobank (**Figure 1**).

**Figure 1.**
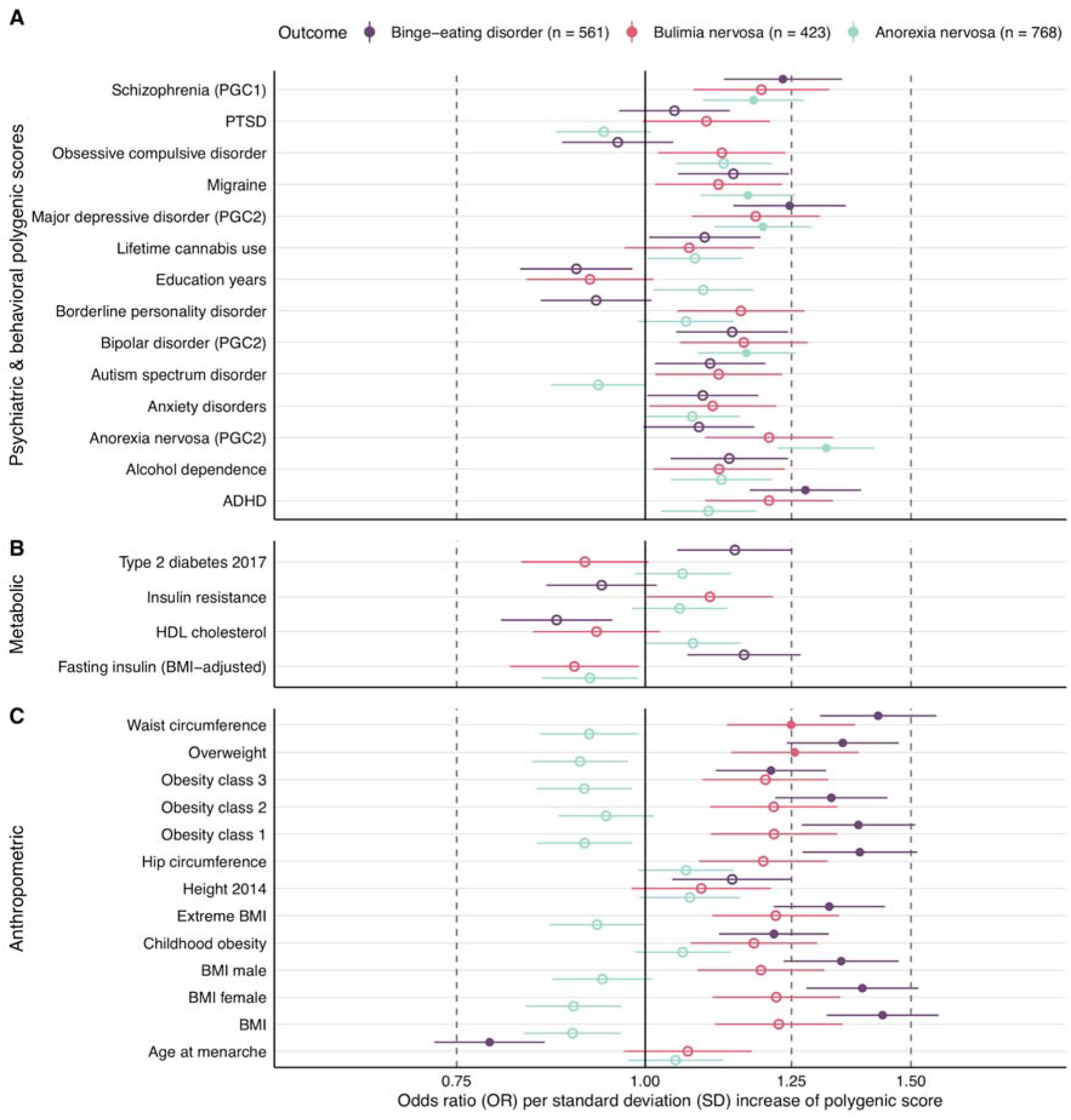
Polygenic scores associated with eating disorders in the UK Biobank. **Panel A** shows psychiatric and behavioral, **Panel B** metabolic, and **Panel C** anthropometric polygenic scores and their association estimates with self-reported or hospital-diagnosed eating disorders in the UK Biobank sample (n = 17,050). Filled dots are statistically significant after adjustment for multiple testing through the false discovery approach. Dots represent odds ratios (ORs) and error bars index 95% confidence intervals obtained via logistic regression and 10,000 permutations to obtain empirical p values.

### Psychiatric disorders and neurological disease

Five polygenic scores reflecting polygenic liability for psychiatric disorders and one neurological disease showed significant associations with eating disorders (**Supplementary Table 4 & 5**). The schizophrenia polygenic score was positively associated with anorexia nervosa (odds ratio [OR] per standard deviation in polygenic score = 1.18, 95% confidence interval [CI]: 1.09, 1.27; *Q* = 0.03) and binge-eating disorder (OR = 1.23, 95% CI: 1.13, 1.35; *Q* = 0.01). The major depressive disorder polygenic score was positively associated with anorexia nervosa (OR = 1.20, 95% CI: 1.11, 1.29; *Q* = 0.01) and binge-eating disorder (OR = 1.25, 95% CI: 1.14, 1.36; *Q* = 0.01). Additionally, the attention deficit hyperactivity disorder polygenic score was positively associated with binge-eating disorder (OR = 1.28, 95% CI: 1.17, 1.39; *Q* = 0.01). The bipolar disorder polygenic score (OR = 1.17, 95% CI: 1.08, 1.26; *Q* = 0.04), the anorexia nervosa polygenic score (OR = 1.32, 95% CI: 1.22, 1.42; *Q* = 0.01), and the migraine polygenic score (OR = 1.17, 95% CI: 1.09, 1.26; *Q* = 0.04) were positively associated with anorexia nervosa.

### Somatic traits

Several polygenic scores indexing polygenic load for anthropometric traits, including waist circumference (OR = 1.43, 95% CI: 1.31, 1.56; *Q* = 0.01), hip circumference (OR = 1.39, 95% CI: 1.27, 1.51; *Q* = 0.01), overweight (OR = 1.35, 95% CI: 1.24, 1.47; *Q* = 0.01), obesity class 1 (OR = 1.38, 95% CI: 1.27; 1.51; *Q* = 0.01), class 2 (OR = 1.33, 95% CI: 1.22; 1.45; *Q* = 0.01) and class 3 (OR = 1.21, 95% CI: 1.11, 1.32; *Q* = 0.04), BMI (OR = 1.44, 95% CI: 1.32; 1.56; *Q* = 0.01) and extreme BMI (OR = 1.32, 95% CI: 1.22, 1.44; *Q* = 0.01) were positively associated with binge-eating disorder. However, it is important to note that these polygenic scores were correlated with each other with *r* = .10 to *r* = .67 (**Supplementary Figure 1**). Additionally, the childhood obesity polygenic score (OR = 1.22, 95% CI: 1.12, 1.33; *Q* = 0.04) was positively associated with binge-eating disorder, while the age at menarche polygenic score (OR = 0.79, 95% CI: 0.72, 0.86; *Q* = 0.01) was negatively associated with binge-eating disorder. Neither the childhood obesity nor the age at menarche polygenic score were associated with anorexia nervosa.

The overweight polygenic score (OR = 1.26, 95% CI: 1.14, 1.38; Q = 0.01) and the waist circumference polygenic score (OR = 1.25, 95% CI: 1.13, 1.38; Q = 0.02) were also positively associated with bulimia nervosa.

### Sex and age differences

The adult obesity polygenic score was more strongly associated with anorexia nervosa than the childhood obesity polygenic score. The difference in the log odds ratios was statistically significant (difference = −0.15, *p* = 0.004). We did not detect age differences in the associations of the obesity polygenic scores with bulimia nervosa or binge-eating disorder (**Supplementary Table 6**). We found no evidence for sex differences in the associations of the body mass index polygenic score with anorexia nervosa, bulimia nervosa, or binge-eating disorder.

### Sensitivity analyses

We compared logistic regression results when assigning participants to all possible eating disorder diagnoses with assigning participants exclusively to one eating disorder diagnosis in the UK Biobank sample. The effect sizes of both analyses correlated with r = 0.98, indicating that the analyses were not sensitive to participant overlap (**Supplementary Figure 2**).

### Independent replication

In ALSPAC (**Supplementary Table 7**), the overweight polygenic score (OR = 1.38, 95% CI: 1.13, 1.69; p_empirical_ = 0.05) and the obesity class 1 polygenic score (OR = 1.23, 95% CI: 1.00; 1.50; p_empirical_ = 0.001) were positively associated with binge-eating disorder. Correlations between these polygenic scores are presented in **Supplementary Figure 3**.

## Discussion

Here, we present the first molecular genetic evidence that the underlying biology differs between binge-type eating disorders and anorexia nervosa and these differences can be captured at the genomic level. We find that polygenic scores for psychiatric disorders and anthropometric traits are associated with binge-type eating disorders (see **Figure 1**). Our UK Biobank analyses propose strong positive associations between bulimia nervosa/binge-eating disorder and anthropometric polygenic scores (e.g., overweight and waist circumference), suggesting that binge eating shares genomic variants with overweight and obesity. In contrast, for anorexia nervosa, the direction of these associations was reversed. Binge-eating disorder also showed an association with the childhood obesity polygenic score while anorexia nervosa did not, further highlighting the genomic differences between binge-type eating disorders and anorexia nervosa. The genetic relationship between binge eating and BMI had been studied previously (Abdulkadir et al., 2020; Bulik et al., 2003) and is replicated by our analyses. It is notable that previous work suggests that genomic variants associated with BMI are predominantly expressed in brain tissue (Finucane et al., 2018). Our results also suggest that these BMI-associated genomic variants are relevant for eating disorders, but may act in opposite directions in binge-type eating disorders and anorexia nervosa.

Using the UK Biobank data, we also found that psychiatric polygenic scores (e.g., schizophrenia and major depressive disorder) were positively associated with both anorexia nervosa and binge-eating disorder (**Figure 1, Panel A**). The ADHD polygenic score was only significantly associated with binge-eating disorder, while the anorexia polygenic score was not associated with bulimia nervosa or binge-eating disorder.

Our findings are in agreement with previous studies that showed genetic correlations between binge eating and depression (Munn-Chernoff et al., 2015) as well as genetic correlations between anorexia nervosa and depression (Wade et al., 2000; Watson et al., 2019), migraine (Mustelin et al., 2014), schizophrenia (Duncan et al., 2017; Watson et al., 2019), bipolar disorder (Stahl et al., 2019) and anthropometric traits (Duncan et al., 2017; Watson et al., 2019). In line with our findings, previous GWASs showed no genomic association between anorexia nervosa and ADHD, whereas an ADHD polygenic score was associated with binge eating in a Swedish twin sample (Capusan et al., 2017; Yao, Kuja-Halkola, et al., 2019). Moreover, a large evidence-base highlights phenotypic overlap between ADHD and binge eating (Cortese et al., 2016; Leventakou et al., 2016; Sonneville et al., 2015).

Large GWASs have recently suggested the involvement of a metabolic component in anorexia nervosa through genetic correlations (Duncan et al., 2017; Watson et al., 2019); however, at the current sample size, we did not observe associations between metabolic polygenic scores and the three primary eating disorders in the UK Biobank most probably due to limited statistical power (**Figure 1, Panel B**).

The association between polygenic liability for higher body mass and eating disorders is further complicated as it shows age dependence (Hübel et al., 2019) and confounding through age at menarche. In both the UK Biobank and the ALSPAC cohort, we found that age at menarche and BMI polygenic scores were negatively correlated with each other. However, the age at menarche polygenic score was only negatively associated with binge-eating disorder but not with anorexia nervosa (**Figure 1, Panel C**). Phenotypic associations between early menarche and bulimia nervosa have previously been reported (Fairburn et al., 1999; Reichborn-Kjennerud et al., 2004), but do not always replicate (Algars et al., 2014). On a genetic level, binge eating correlated negatively with age at menarche in a twin study (Baker et al., 2012) and anorexia nervosa was not genetically correlated with age at menarche in our polygenic score analysis nor in previous GWASs (Duncan et al., 2017; Watson et al., 2019). This suggests that the association between age at menarche and eating disorders may be driven by symptoms related to binge eating rather than restriction.

Furthermore, ADHD and BMI show a positive genetic correlation (Demontis et al., 2019; Hübel et al., 2019) and in our analysis the ADHD polygenic score was associated with binge-eating disorder. Clinically, the ADHD medication lisdexamfetamine is used to treat binge-eating disorder (Hudson et al., 2017). This combined evidence suggests that the shared biology between ADHD and BMI may contribute to binge eating. These novel findings warrant further research; investigating how genomic variants associated with ADHD, commonly characterized by inattention and impulsivity, are also associated with binge-eating disorder and contribute to the high comorbidity between ADHD and obesity (Cortese et al., 2016; Leventakou et al., 2016; Sonneville et al., 2015).

Our findings must be interpreted in the light of the following limitations: the number of individuals with eating disorders in both samples was relatively low, some of the eating disorder diagnoses and symptoms were self-reported, and the samples only included white British participants. Furthermore, we were unable to differentiate between binge-eating/purging and restricting anorexia nervosa as subtype information was not available. However, symptoms and derived diagnoses in the ALSPAC cohort were obtained both from parents and adolescents, representing the gold standard in child and adolescent psychiatry, strengthening the validity of the diagnoses. Further, replication studies are essential to lend confidence to the results reported here.

Our findings show for the first time that similarities exist in the genomic psychiatric liability for binge-type eating disorders and anorexia nervosa. However, we find clear differences between binge-type eating disorders and anorexia nervosa in the underlying biology of body mass regulation at the genomic level. These findings open important avenues for translational research relevant to eating disorder phenotypes and overlap between eating disorders. Specifically, it remains unknown which biological mechanisms are shared between ADHD and binge eating.

## Data Availability

This study is based on data from the UK Biobank. Researchers can apply for the data via the following website: https://www.ukbiobank.ac.uk/. This study is based on data from the ALSPAC study (http://www.bristol.ac.uk/alspac/). Interested researchers can apply for data access with the University of Bristol, UK. Analysis scripts can be requested from the authors.

## Acknowledgements

We are deeply grateful to all the families who took part in this study, the midwives for their help in recruiting them, and the whole ALSPAC team, which includes interviewers, computer and laboratory technicians, clerical workers, research scientists, volunteers, managers, receptionists and nurses. This study was completed as part of approved UK Biobank study applications 27546 to Prof Breen.

## Funding

This study represents independent research part funded by the UK National Institute for Health Research (NIHR) Biomedical Research Centre at South London and Maudsley NHS Foundation Trust and King’s College London. The views expressed are those of the author(s) and not necessarily those of the UK NHS, the NIHR or the Department of Health. High performance computing facilities were funded with capital equipment grants from the GSTT Charity (TR130505) and Maudsley Charity (980). This work was supported by the UK Medical Research Council and the Medical Research Foundation (ref: MR/R004803/1). The UK Medical Research Council and Wellcome (Grant ref: 102215/2/13/2 and 217065/Z/19/Z) and the University of Bristol provide core support for ALSPAC. A comprehensive list of grants funding is available on the ALSPAC website (http://www.bristol.ac.uk/alspac/external/documents/grant-acknowledgements.pdf); This research was specifically funded by the NIHR (CS/01/2008/014), the NIH (MH087786-01). GWAS data was generated by Sample Logistics and Genotyping Facilities at Wellcome Sanger Institute and LabCorp (Laboratory Corporation of America) using support from 23andMe. NM and CB acknowledge funding from the National Institute of Mental Health (R21 MH115397). CB acknowledges funding from the Swedish Research Council (VR Dnr: 538-2013-8864), the National Institute of Mental Health (R21MH115397; R01 MH109528; R01MH120170; R01MH119084; R01MH120170), and Lundbeckfonden. MH is supported by fellowship from the Medical Research Council UK (MR/T027843/1). The content is solely the responsibility of the authors and does not necessarily represent the official views of the National Institutes of Health. The funders were not involved in the design or conduct of the study; collection, management, analysis, or interpretation of the data; or preparation, review, or approval of the manuscript.

## Declaration of interest

Dr. Breen has received grant funding from and served as a consultant to Eli Lilly, has received honoraria from Illumina and has served on advisory boards for Otsuka. Dr. Bulik is a grant recipient from and has served on advisory boards for Shire and is a consultant for Idorsia. She receives royalties from Pearson. She is a grant recipient from Lundbeckfonden. All other authors have indicated they have no conflicts of interest to disclose.

## Author Contribution

CH, MA, MH analysed the data. CH, MA, MH drafted the manuscript. CMB, NM, RFL, and GB supervised the work. All authors substantially contributed to the conception and interpretation of the work, revised the manuscript for important intellectual content and approved the final version. All authors agree to be accountable for all aspects of this work.

## Notes

### Summary of Updates

We have split the manuscript in 3 separate manuscripts because it was difficult to follow.

