## Supplementary Info for "One size does not fit all. Genomics differentiates among binge-eating disorder, bulimia nervosa, and anorexia nervosa"

### Genotyping, imputation and quality control in the UK Biobank

### Genotyping, imputation and quality control in the Avon Longitudinal Study of Parents and Children (ALSPAC)

**Supplementary Figure 1.** Heatmap of the correlations across polygenic scores calculated in the UK Biobank subsample (n = 17,035).

**Supplementary Figure 2.** Sensitivity analysis comparing logistic regression results when assigning participants to all possible eating disorder diagnoses or assigning participants exclusively to one eating disorder diagnosis in the UK Biobank sample (n = 17,035).

#### **Correlations between polygenic scores in ALSPAC**

**Supplementary Figure 3.** Heatmap of the correlations across polygenic scores calculated in the ALSPAC subsample (n = 4,573).

### Genotyping, imputation and quality control in the UK Biobank

Blood samples were genotyped on two arrays, which share nearly all of their content: the UKBileve array (*N* = 49,949) or the UK Biobank Axiom array (*N* = 438,414). Genotyping was conducted by Affymetrix and was distributed across 33 different batches of approximately 4,700 samples. UK Biobank provides extensive information on sample processing on its web site, <http://biobank.ctsu.ox.ac.uk/crystal/refer.cgi?id=155583>, and details of the Axiom array are available at <http://media.affymetrix.com/support/downloads/manuals/axiom_2_assay_auto_workflow_user_guide.pdf>. UK Biobank performed stringent quality control on the genotyping data at the Wellcome Trust Centre for Human Genetics (WTCHG). For further details, see: <http://biobank.ctsu.ox.ac.uk/crystal/refer.cgi?id=155580>. Prior to imputation, all variant sites with a call rate below 90% were filtered out. Imputation was carried out by UK Biobank using the IMPUTE3 program and a merged UK10K-1000 Genomes Phase 3 reference panel (details available at [http://biobank.ctsu.ox.ac.uk/crystal/refer.cgi?id=157020)](http://biobank.ctsu.ox.ac.uk/crystal/refer.cgi?id=157020).

We applied additional quality controls. Specifically, we excluded genotyped participants who were pregnant (*N* = 105) or non-European participants identified by k-means clustering (*k* = 4) on the first two principal components derived from the genotype data, and we excluded related individuals (KING relatedness metric >0.088, equivalent to a relatedness value of 0.25; *N* = 7,765), or participants with biological sex mismatch.

SNPs were excluded if they had a minor allele frequency (MAF) smaller than 1%, if they deviated substantially from Hardy-Weinberg equilibrium (HWE test, *p*<10^-7^), SNP missingness > 0.02, or individual missingness > 0.02 [[1]](https://paperpile.com/c/oL9DfW/G1LXC). This left a total of 560,178 SNPs and 385,753 for analysis.

### Genotyping, imputation and quality control in the Avon Longitudinal Study of Parents and Children (ALSPAC)

Genotype data were available for 9,915 children out of the total of 15,247 ALSPAC participants. Participants were genome-wide genotyped on the Illumina HumanHap550 quad chip. Individuals with disproportionate levels of individual missingness (i.e., >3%), insufficient sample replication (identity by descent < 0.1), biological sex mismatch, and non-European ancestry (as defined by multi-dimensional scaling using the HapMap Phase II, release 22, reference populations) were excluded. SNPs with a minor allele frequency (MAF) of < 1%, excessive missingness (i.e., call rate < 95%), or a departure from the Hardy–Weinberg equilibrium (P value < 5 x 10-7) were removed. Imputation was conducted with Impute3 using the HRC 1.0 as the reference panel [[1]](https://paperpile.com/c/oL9DfW/G1LXC) and phasing was carried out using ShapeIT (v2.r644). Finally, post-imputation quality control checks were performed; any SNPs with MAF less than 1%, Impute3 information quality metric of < 0.8, and not confirming to Hardy-Weinberg equilibrium (P < 5 × 10-7) were removed. After data cleaning, a total of 8,654 individuals (4,225 females and 4,429 males) and 4,054,653 SNPs remained eligible for analyses.

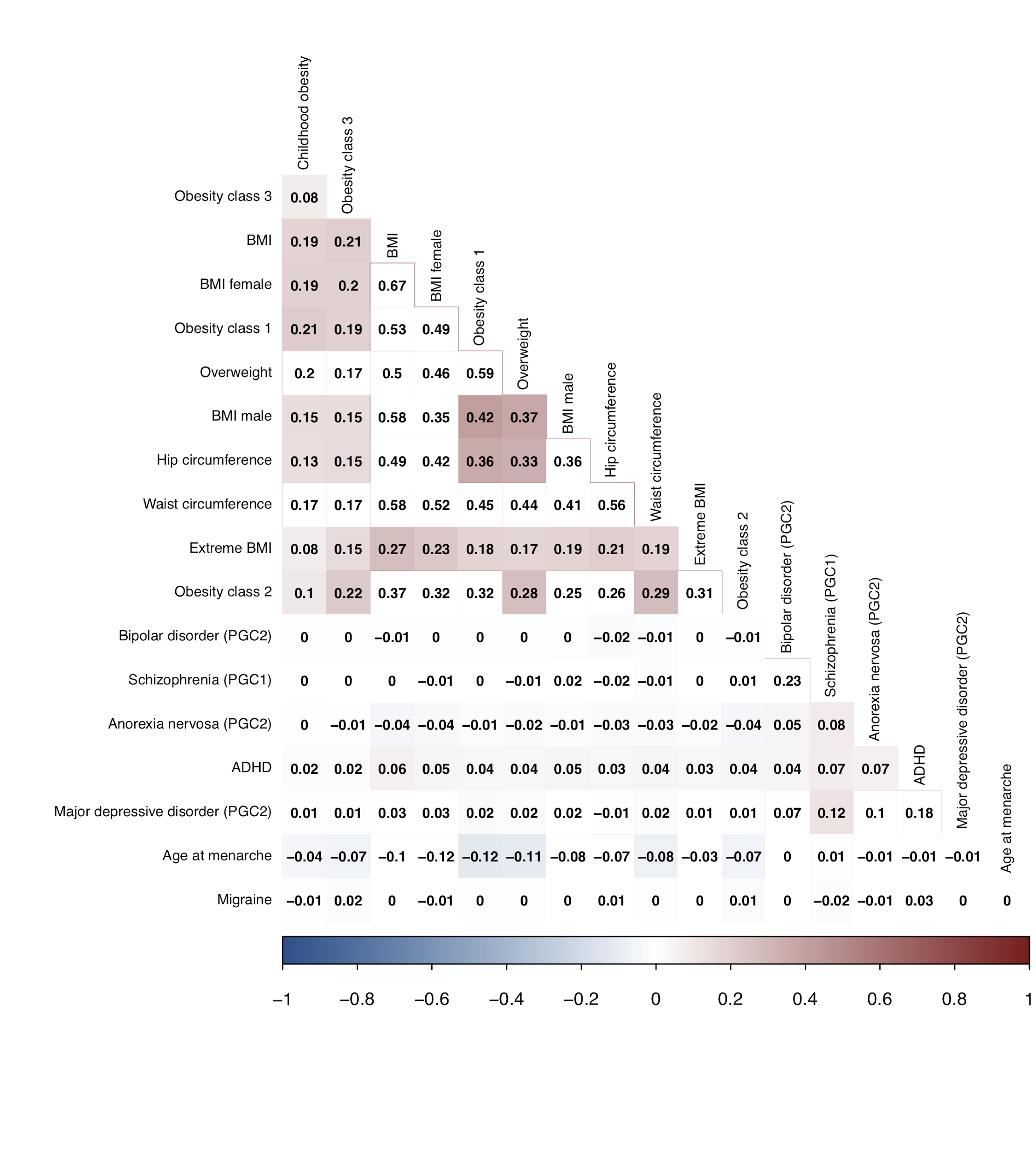

***Supplementary Figure 1.*** *Heatmap of the correlations across polygenic scores calculated in the UK Biobank subsample (n = 17,035). We calculated Pearson correlation coefficients as the polygenic scores were normally distributed in our subsample. Coloured fields are significant with a p value less than 0.01. Red indexes positive correlations and blue negative correlations. Correlations shown in white squares were not significant. We used hierarchical clustering to display the correlations.*

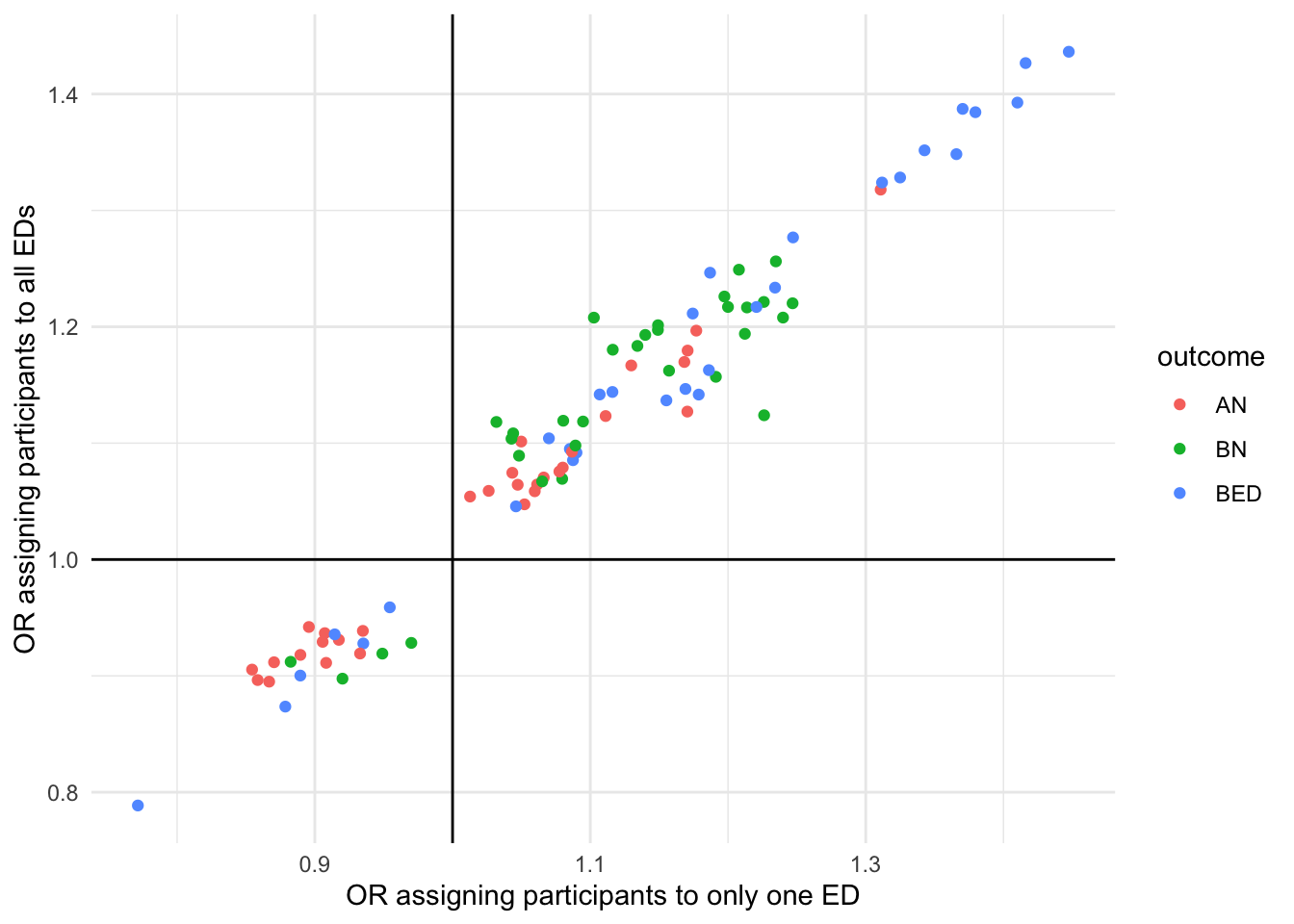

***Supplementary Figure 2.*** *Sensitivity analysis comparing logistic regression results when assigning participants to all possible eating disorder diagnoses or assigning participants exclusively to one eating disorder diagnosis in the UK Biobank sample (n = 17,035).*

#### **Correlations between polygenic scores in ALSPAC**

The anthropometric polygenic scores showed significant intercorrelations, ranging between *r* = .06 and *r* = .64 (**Supplementary Figure 3**). Because of these properties we only report results on the overweight polygenic score in the results section. Additionally, the childhood obesity polygenic score (*r* = ~.10) was positively correlated and the age at menarche polygenic score was negatively correlated (*r* = ~-.10) with other anthropometric polygenic scores.

The schizophrenia polygenic score correlated with the ADHD (*r* = .05), the major depressive disorder (*r* = . 13), and the anorexia nervosa polygenic score (*r* = .10). The major depressive disorder polygenic score additionally correlated with polygenic scores of anorexia nervosa (*r* = .16) and ADHD (r = .21), indicating that individuals can carry genetic liability for different psychiatric disorders concurrently.

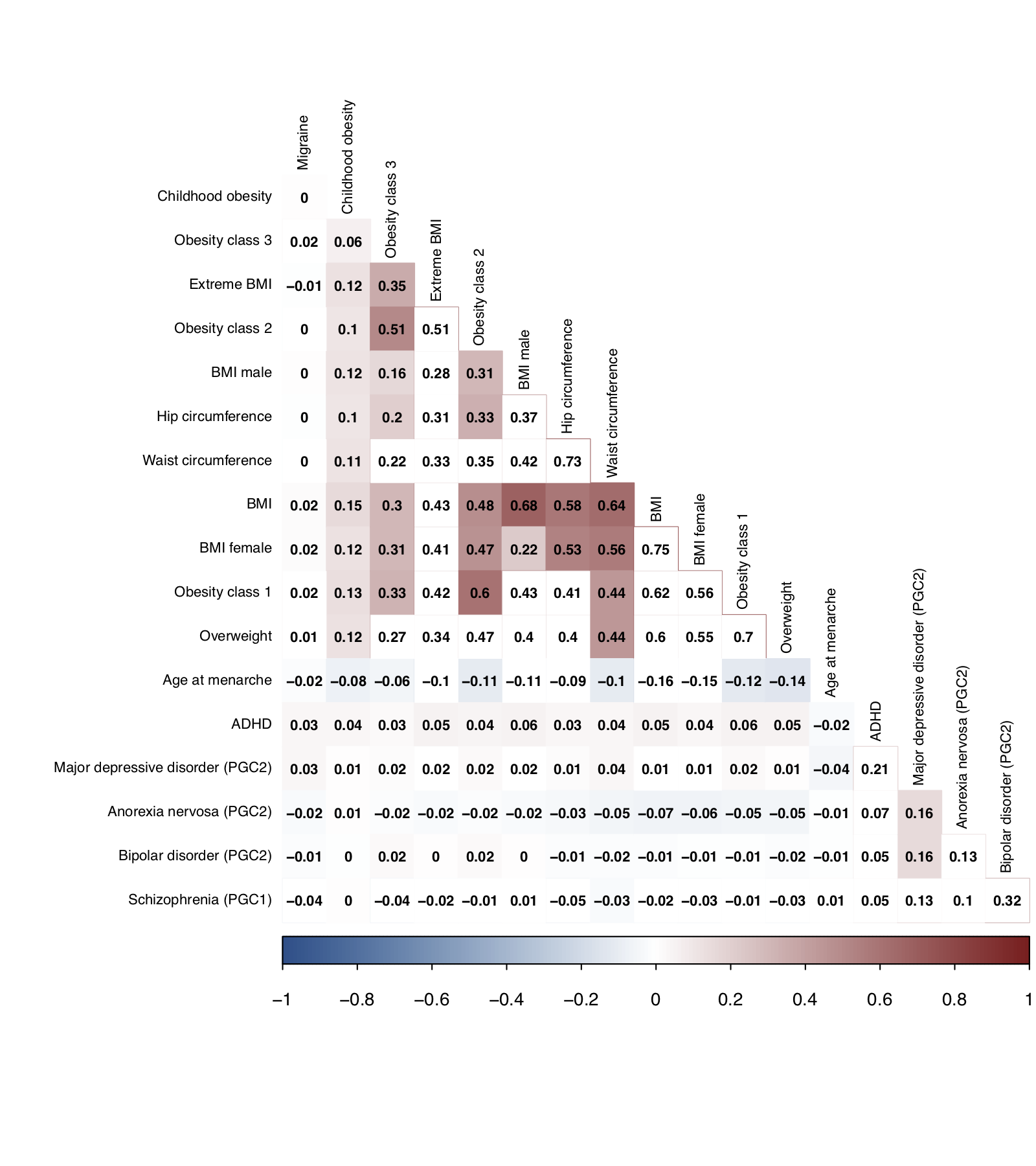

***Supplementary Figure 3.*** *Heatmap of the correlations across polygenic scores calculated in the ALSPAC subsample (n = 4,573). We calculated Pearson correlation coefficients as the polygenic scores were normally distributed in our subsample. Coloured fields are significant with a p value less than 0.01. Red indexes positive correlations and blue negative correlations. Correlations shown in white squares were not significant. We used hierarchical clustering to display the correlations.*
